# Maternal ADHD and exclusive breastfeeding: A prospective community-based cohort study

**DOI:** 10.64898/2026.11.02.26367066

**Authors:** Tatiana Dessy, Cindy-Lee Dennis

**Author notes:** **Corresponding Author** Tatiana Dessy, PhD, Lunenfeld-Tanenbaum Research Institute, 600 University Ave, Toronto, ON, Canada, M5G 1X5.

## Abstract

**Importance:** Maternal attention-deficit disorder with or without hyperactivity (ADHD) is an established risk factor for adverse perinatal outcomes that remains understudied to breastfeeding.

**Objective:** To estimate the effect of maternal ADHD on 6-month exclusive breastfeeding.

**Design:** A prospective longitudinal community-based cohort study.

**Setting:** Canada, 2014-2020.

**Participants:** We included 3217 English-literate mother-father dyads with internet or telephone access, comprising women with singleton birth within 3 weeks postpartum (study baseline) and were cohabiting with the biological father. Women with major psychiatric conditions or suicidality were excluded. All participants provided informed consent at recruitment. Data were collected online through self-report questionnaires.

**Exposure:** Maternal ADHD was measured at baseline using the Adult ADHD Self-Report Scale (sum of scores ≥14).

**Main Outcomes and Measures:** We defined 6-month exclusive breastfeeding based on maternal reported infant feeding, assessed at 3 and 6 months. Maternal childhood adversity (Adverse Childhood Experiences International Questionnaire, ACE-IQ; sum score) was included as a confounder. The average treatment effect on the treated (ATT) was estimated using linear probability models. We calculated 95% percentile bootstrap CI. Missing data were handled using multiple imputation by chained equations with 10 iterations.

**Results:** Complete-case analyses included 2071/3217 mother-father dyads, most of which were Canadian-born (79%). Childhood adversity, mental health conditions, and substance use were higher among mothers who screened positive for ADHD (4%) than among those who did not: the median number of ACE was 5 IQR 4-6; 38% self-reported prenatal depression, 13% smoked cigarettes and 19% drank alcohol during pregnancy; 11% of infants used the neonatal intensive care unit. Notably, though 90% of mothers with ADHD intended to breastfeed exclusively only 28% breastfed exclusively for 6 months. Among families with maternal ADHD, the effect of maternal ADHD on 6-month exclusive breastfeeding was ATT −0.105, 95% CI −0.209 to −0.003 in the model adjusted for maternal ACE and paternal ADHD. With multiple imputation: ATT −0.109, 95% CI −0.213 to −0.004.

**Conclusions and Relevance:** Mothers with ADHD intend to breastfeed their infants exclusively, but additional challenges shorten the duration of exclusive breastfeeding, indicating a need for interventions tailored to neurodevelopmental differences.

**KEY POINTS:** *Question:* Does maternal ADHD influence exclusive breastfeeding duration?

*Findings:* In this community-based cohort study of 3217 mother-father dyads in Canada, most mothers with ADHD intended to breastfeed exclusively, but few actually maintained it for 6 months. Maternal ADHD predicted lower probability of exclusive breastfeeding until 6 months among families with maternal ADHD.

*Meaning:* To equitably promote maternal and infant well-being, early detection and support for breastfeeding challenges are needed for mothers with ADHD.

## INTRODUCTION

Attention-deficit disorder with or without hyperactivity (ADHD) is a neurodevelopmental condition characterized by executive dysfunction, emotional dysregulation, and heightened susceptibility to stressors [1]. ADHD affects about 2-3% of adults, with a diagnosis-based prevalence of 0.11% among women who recently gave birth in the United States [2].

Exclusive breastfeeding is an important pediatric health practice, but maintaining it can be challenging, even among women who intend to breastfeed [3]. Maternal ADHD may influence breastfeeding experiences through executive functioning, emotional regulation, and other postpartum challenges, yet direct evidence remains limited. With maternal emotional dysregulation, mental health illness, and low social support, breastfeeding becomes increasingly difficult [3–6]. However 6-month exclusive breastfeeding is a therapeutic feeding target, fostering infant development through immune and relational pathways [4,7]. In a 2000-2014 Swedish national registry-based birth cohort, parental ADHD was associated with an increased rate of child assault or victimization (HR 1.82, 95% CI 1.66-1.99) [8]. Both maternal and paternal ADHD are associated with infant well-being, yet remain understudied as upstream determinants of exclusive breastfeeding duration [9].

In Canada and other high-income settings, targeted preventive interventions to promote infant development through breastfeeding support are available to families of lower socioeconomic status [10,11]. However few (if any) interventions are tailored to families with parental ADHD, leaving these infants without adequate developmental support [12–14]. First, prospective evidence is needed to determine whether maternal ADHD directly influences exclusive breastfeeding, accounting for relevant maternal and family characteristics.

In this study, we aim to estimate the effect of maternal ADHD on exclusive breastfeeding up to 6 months within a causal inference framework using observational data from the IMPACT prospective cohort study (2014-2020, Canada) [15]. We hypothesize that common breastfeeding challenges – including limited access to adequate lactation support – compounded by ADHD symptoms such as inattentiveness and emotional dysregulation inhibit exclusive breastfeeding.

## METHODS

### Study Design

We conducted a prospective longitudinal cohort study within the ‘**I**mpact of **M**aternal and **P**aternal mental health: **A**ssessing **C**oncurrent depression, anxiety, and comorbidity in **T**he Canadian family’ (IMPACT) study [15]. We followed the updated Strengthening the Reporting of Observational Studies in Epidemiology (STROBE) reporting guidelines for this study [16].

### Setting

The IMPACT study was based at Unity Health Toronto with recruitment in the Greater Toronto Area (in-hospital and via community outreach) and across Canada (via social media). Recruitment started [Month Day], 2014 (ending [Month Day], 2018) and data collection ended [Month Day], 2020.

### Participants

Women who recently gave birth to a single child (<3 weeks) at >33 weeks gestation and were cohabiting with the infant’s biological father were included. Both parents needed to be literate in English and have internet or telephone access to participate. Participants (women) were excluded for ethical reasons if they had major psychiatric conditions – either diagnosed schizophrenia, bipolar disorder, or psychosis – or women experiencing symptoms of suicidality.

### Ethical Approval

The IMPACT study received ethical approval from the University of Toronto Research Ethics Board (#29655) and from participating community-based hospitals in the Greater Toronto Area. All participants provided informed consent before study inception.

### Exposure

The main exposure was maternal ADHD. We measured core ADHD symptoms (ie, inattentiveness, hyperactivity, and impulsivity) at baseline using the psychometrically validated Adult ADHD Self-Report Scale (sum score range 0-24 for items from Part A of the questionnaire) [17]. This scale was validated to screen for clinically diagnosed ADHD; a sum score ≥14 signifies a ‘positive’ ADHD result [18].

### Outcomes and Measures

#### Exclusive Breastfeeding

The main outcome was 6-month exclusive breastfeeding. To calculate this variable, we used maternal reports of infant feeding over the past 7 days, assessed at 3 and 6 months postpartum. The question ‘*Please indicate how you are breastfeeding your baby*’ included a response option ‘*Exclusive breastfeeding - My baby has received only breast milk in the past 7 days (may include breast milk in a bottle or vitamins)*’ which identified exclusive breastfeeding. Exclusive breastfeeding at both 3 and 6 months postpartum indicated “6-month exclusive breastfeeding” otherwise, we considered that mixed feeding or non-breastmilk was used during the 6-month period.

#### Confounding

Measured confounders of the relationship between maternal ADHD and 6-month exclusive breastfeeding included only childhood adversity: given that ADHD is a heritable neurodevelopmental condition present from birth, confounders are hypothesized to be either genetic or, for ADHD severity, environmental [19,20], which we capture via childhood adversity. We measured childhood adversity using the psychometrically validated Adverse Childhood Experiences International Questionnaire (ACE-IQ) [21]. We calculated the number of ACE as a sum score across 13 categories including: neglect (emotional, physical); abuse (emotional, physical); violence against a household member; living with a household member who was a substance abuser, mentally ill or suicidal, incarcerated or jailed; bullying; having a precarious family composition (ie, single parent, orphan, parental separation or divorce); community violence; and collective violence.

#### Covariates

Other baseline measures included Canadian birth and newcomer status (ie, having lived in Canada for <5 years); unemployment; substance use during pregnancy (cigarettes, alcohol, other); infant sex at birth; preterm birth (gestational age <37 weeks); low birth weight (birth weight <2500 g); admission to the neonatal intensive care unit (NICU); self-reported intention to breastfeed exclusively; and self-reported depression and anxiety during pregnancy assessed via yes/no items.

We also calculated maternal reported intimate partner violence (IPV) at 3 months using the Woman Abuse Screening Tool, short version, which has high sensitivity but low specificity [22]. Any positive response to both of the first two items on relationship tension indicated potential abuse.

### Statistical Analysis

Participants’ baseline characteristics and 3- and 6-month exclusive breastfeeding were described by exposure group (ie, maternal ADHD vs none) using absolute and relative frequencies or median with interquartile range (IQR). To assess selection bias due to attrition, we compared baseline characteristics of participants with complete data (ie, those with data for maternal ADHD, paternal ADHD, 6-month exclusive breastfeeding, and maternal number of ACE) to those with missing data. We calculated standardized mean differences (SMD) and used SMD >.2 as a significance cut-off.

#### Causal Effect of Maternal ADHD on 6-month Exclusive Breastfeeding

We estimated linear probability models of 6-month exclusive breastfeeding, adjusting for maternal ADHD and the number of ACE. First, we used complete-cases only. The average treatment on the treated (ATT) for 6-month exclusive breastfeeding was calculated for maternal ADHD among families with maternal ADHD. The ATT was defined as the regression coefficient for ADHD in a heterogeneous-effects model [23,24] (eMethods1). In sensitivity analyses, we adjusted the linear probability model of 6-month exclusive breastfeeding for paternal ADHD (ASRS sum score). We estimated the association between paternal ADHD and maternal reports of IPV at 3 months using Cohen’s *d* and linear probability regression models. We also repeated the analyses using 3-month exclusive breastfeeding as a secondary outcome. Statistical inference on the ATT was based on heteroskedasticity-robust standard errors and 95% bootstrap percentile CI (B=3000 replications, with replacement). Second, to account for attrition bias, we used inverse probability of attrition weighting to estimate the ATT [25]. Attrition probabilities were calculated using baseline covariates which differed in distribution or proportion between included and excluded participants. Attrition weights were calculated and standard stabilization methods were used as needed. Third, to improve statistical precision of the ATT, we used multiply imputed data by chained equations (m=10 iterations) within R package ‘mice’ [26]. All statistical analyses were conducted in R version 4.6.0 [27].

## RESULTS

### Study Participants

Our analytic sample included 2071/3217 (64%) of mother-father dyads with complete data on relevant variables (Figure 1, Table 1). Maternal ADHD was identified in 4% of mothers reporting ASRS sum scores ≥14. On average, paternal ADHD among families with maternal ADHD was not clinically significant: mean ASRS score 8.9 SD 3.8, with 8% reporting ASRS sum scores ≥14. Most participants were Canadian-born and among those born outside of Canada, up to a fourth were newcomers. Most participants were employed full-time during pregnancy. Maternal childhood adversity differed significantly between exposure groups, with a higher median ACE among mothers with ADHD than among those without. Paternal childhood adversity was similar between groups. Cigarette and alcohol use were higher among mothers with ADHD than among those without. Self-reported depression and anxiety during pregnancy were more common among mothers with ADHD than among those without. There were no major differences in infant sex at birth between exposure groups; preterm birth and low birth weight were rare, and NICU use was higher among infants born to mothers with ADHD. Most mothers intended to breastfeed exclusively, but fewer did so continuously until 6 months among mothers with ADHD compared to those without ADHD. Excluded participants (n=1146) were less intent on exclusive breastfeeding (eTable 1).

**Table 1.** Study participants’ characteristics by maternal ADHD <3 weeks postpartum (IMPACT, 2014-2020, Canada).

|  | **Maternal ADHD<sup>a</sup>, <3 weeks postpartum** |  |  |  |
|---|---|---|---|---|
|  | **Yes** |  | **No** |  |
|  | **Mothers (n=130)** | **Fathers (n=130)** | **Mothers (n=3077)** | **Fathers (n=3077)** |
| **Socio-demographics** |  |  |  |  |
| Canadian born, No. (%) | 110 (84.6) | 102 (78.5) | 2418 (78.7) | 2315 (75.4) |
| Newcomer, No. (%) | 4 (20.0) | 7 (25.0) | 133 (20.7) | 162 (22.0) |
| Unemployed, No. (%) | 17 (13.1) | <sup>†</sup> | 305 (9.9) | <sup>†</sup> |
| Childhood adversity, number of ACE<sup>b</sup>, median [IQR<sup>c</sup>] | 5.0 [4.0, 6.3] | 4.0 [3.0, 6.0] | 3.0 [2.0, 5.0] | 4.0 [2.0, 5.0] |
| **Substance use, pregnancy** |  |  |  |  |
| Cigarettes, No. (%) | 17 (13.1) | 25 (19.2) | <sup>†</sup> | 355 (11.5) |
| Alcohol, No. (%) | 24 (18.5) | 95 (73.1) | 460 (15.0) | 2340 (76.1) |
| Other substances, No. (%) | <sup>†</sup> | 18 (13.8) | <sup>†</sup> | 219 (7.1) |
| **Mental health, pregnancy** |  |  |  |  |
| Depression, No. (%) | 49 (37.7) | 12 (9.2) | 263 (8.5) | <sup>†</sup> |
| Anxiety, No. (%) | 51 (39.2) | 12 (9.2) | 443 (14.4) | 312 (10.1) |
| **Infant characteristics** |  |  |  |  |
| Sex at birth, male, No. (%) | 65 (50.0) |  | 1579 (51.3) |  |
| Preterm birth, No. (%) | <sup>†</sup> |  | <sup>†</sup> |  |
| Low birth weight, No. (%) | <sup>†</sup> |  | <sup>†</sup> |  |
| NICU<sup>d</sup> use, No. (%) | 15 (11.5) |  | 267 (8.7) |  |
| **Exclusive breastfeeding** |  |  |  |  |
| Intention, No. (%) | 116 (89.2) | 88 (72.1) | 2806 (91.3) | 2078 (73.6) |
| Duration, 3 months, No. (%) | 78 (75.0) |  | 2059 (79.8) |  |
| Duration, 6 months, No. (%) | 21 (27.6) |  | 759 (37.5) |  |

**<sup>a</sup>: Mothers’ responses to the Adult ADHD Self-Report Scale [17].**
**<sup>b</sup>: Adverse Childhood Experiences International Questionnaire (ACE-IQ) [21].**
**<sup>c</sup>: Interquartile range, IQR.**
**<sup>d</sup>: Neonatal Intensive Care Unit, NICU.**
**<sup>†</sup>: Relative frequency <5%.**

**Figure 1.**
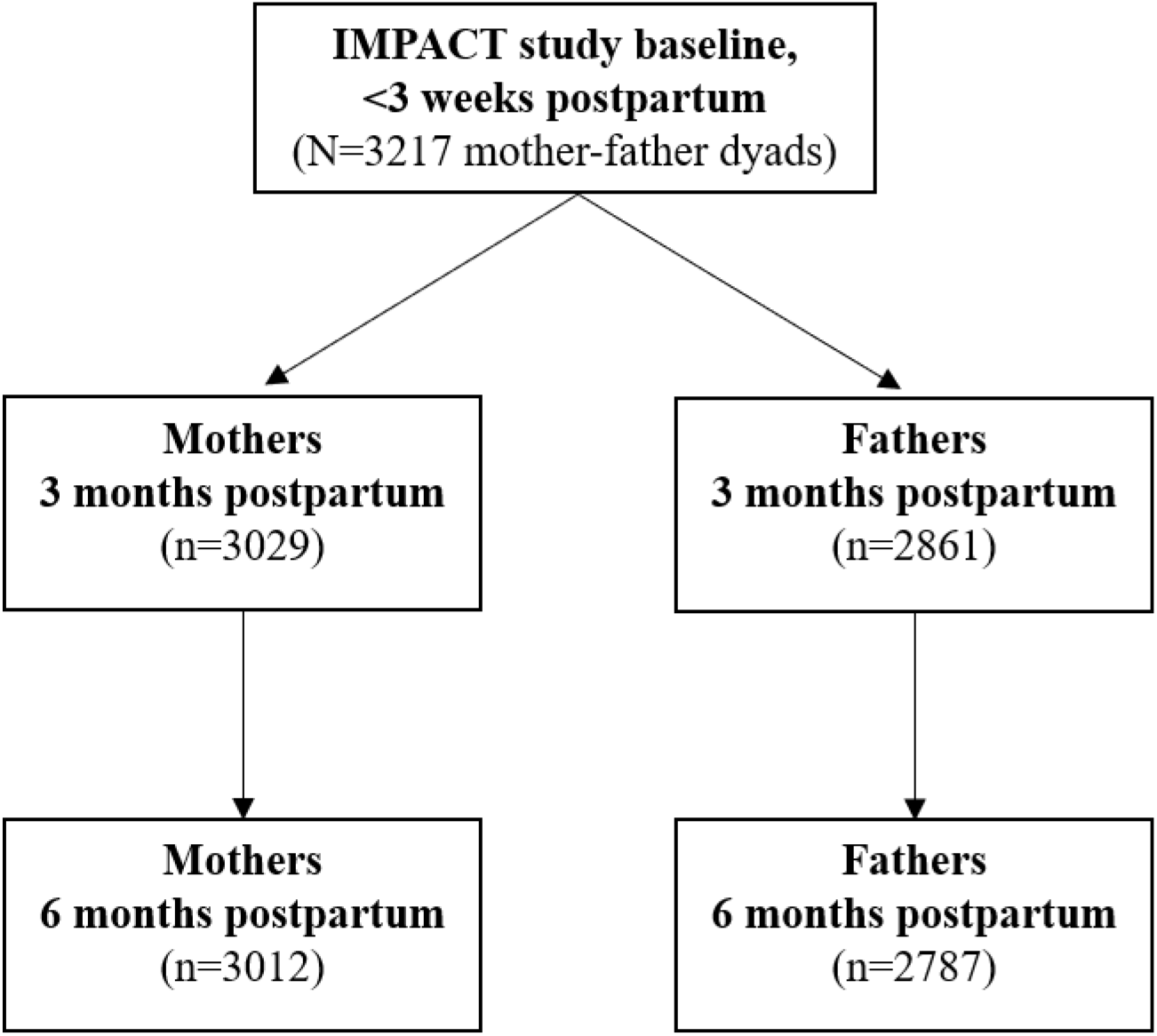
Flow chart (IMPACT, 2014-2020, Canada). Adapted from [15].

### Maternal ADHD

Higher maternal ADHD scores were associated with a lower probability of exclusive breastfeeding, after accounting for maternal childhood adversity and paternal ADHD scores (*β* - 0.006, SE 0.003, 95% CI −0.013 to −0.000, *P*=.04). Estimated effect sizes of ADHD (ASRS scores ≥14) on 6-month exclusive breastfeeding were small. Using maternal ADHD, we observed 28% vs. 37% of mothers reporting 6-month exclusive breastfeeding among families with vs. without maternal ADHD, respectively: Cohen *d* 0.19, 95% CI −0.04 to 0.42. Using paternal ADHD, we observed 43% vs. 36% of mothers reporting 6-month exclusive breastfeeding among families with vs. without paternal ADHD, respectively: Cohen *d* −0.14, 95% CI −0.30 to 0.02.

Among families with maternal ADHD (4%), the complete-case ATT was −0.105, SE 0.053, with 95% CI −0.209 to 0.005 in the model adjusted for maternal childhood adversity, whereas the ATT was significantly smaller in the unadjusted model (Table 2). In complete-case analyses using the model augmented with paternal ADHD, the effect of maternal ADHD on 6-month exclusive breastfeeding was ATT −0.105, SE 0.053, with 95% CI −0.209 to −0.003. (For 3-month exclusive breastfeeding, we estimated ATT −0.013, 95% CI −0.056 to 0.016.)

**Table 2.** Effect of maternal ADHD <3 weeks postpartum on 6-month exclusive breastfeeding among families with maternal ADHD.

| **Model** | **6-month exclusive breastfeeding** |  |  |
|---|---|---|---|
|  | **ATT<sup>a</sup>** | **SE<sup>b</sup>** | **95% CI<sup>c</sup>** |
| **Complete-cases** |  |  |  |
| Unadjusted | -0.093 | 0.053 | (-0.198 to 0.013) |
| Adjusted<sup>d</sup> | -0.105 | 0.053 | (-0.209 to 0.005) |
| Adjusted<sup>e</sup> | -0.105 | 0.053 | (-0.209 to -0.003) |
| **IPAW<sup>f</sup>** |  |  |  |
| Adjusted<sup>e</sup> | -0.107 | 0.060 | (-0.221 to 0.014) |
| **Multiple imputation<sup>g</sup>** |  |  |  |
| Adjusted<sup>e</sup> | -0.109 | 0.053 | (-0.213 to -0.004) |

**<sup>a</sup>: Average treatment effect on the treated, ATT.**
**<sup>b</sup>: Heteroskedasticity-robust SE.**
**<sup>c</sup>: Bootstrap percentile CI for complete-case and IPAW; CI assuming normality of estimates for multiple imputation.**
**<sup>d</sup>: Maternal childhood adversity.**
**<sup>e</sup>: Maternal childhood adversity and paternal ADHD.**
**<sup>f</sup>: Inverse probability of attrition weighting [25]. The attrition model was adjusted for baseline maternal and paternal intention to breastfeed and Canadian birth.**
**<sup>g</sup>: Multiple imputation by chained equations (m=10 iterations). The imputation model included: intention to breastfeed exclusively, infant sex at birth, neonatal intensive care unit use, Canadian birth, unemployment, prenatal substance use, prenatal depression, prenatal anxiety, ADHD, ACE, and exclusive breastfeeding.**

### Considering Intimate Partner Violence

The prevalence of maternal reported IPV at 3 months was 40%. Using paternal ADHD (ASRS um scores ≥14) we observed 54% vs 39% of mothers reporting IPV among families with vs. without paternal ADHD: Cohen *d* −0.30, 95% CI −0.43 to −0.16 for IPV. In univariate linear probability models of IPV, we observed higher odds of IPV with higher paternal ADHD scores: *β* 0.013, SE 0.002, *P*<.001. In complete-case analyses, the ATT of maternal ADHD on 6-month exclusive breastfeeding in the model augmented with paternal ADHD scores and IPV was: ATT −0.11, 95% CI −0.2211 to −0.0004.

## DISCUSSION

### Main Findings

ADHD is the most common neurodevelopmental disorder among pregnant women. In this prospective, community-based cohort study of 3217 mother-father dyads in Canada, we observed that intention to breastfeed exclusively was similar among mothers with and without ADHD. However among mothers with ADHD, maternal ADHD was associated with lower rates of exclusive breastfeeding at 6 months. To our knowledge, this study is the first to estimate the causal effect of maternal ADHD on exclusive breastfeeding at 6 months. Moreover, our study considered the family unit, including paternal ADHD as a potential contributor to 6-month exclusive breastfeeding. Overall, we proposed a framework to identify an ATT for parental ADHD, an understudied upstream determinant of perinatal mental health (eg, symptoms of depression) and infant development (eg, immunity).

### Comparison With Prior Literature

We observed high proportions of exclusive breastfeeding intention, consistent with findings on intention to breastfeed among new mothers in other Canadian settings [28]. In the cross-sectional study of 944 new mothers by Delawarde-Saïas et al (2021; Quebec, Canada) 94% of participants reported wanting to breastfeed their child, whether exclusively or not [28]. We observed that approximately 90% of participants intended to breastfeed exclusively, indicating strong motivation to meet recommended therapeutic targets [7].

Prior literature on the putative influence of maternal ADHD on 6-month exclusive breastfeeding is limited to a small prospective cohort study with important methodological limitations [9]. In their 2019 study of 164 mother-infant dyads in Israel, Freund-Azaria et al observed a negative association between ADHD measured using maternal ASRS scores and 6-month exclusive breastfeeding. In this study, we reach the same conclusion within a formal causal inference framework, with a larger sample, and considering both parents.

We included paternal ADHD as a potential risk factor contributing to exclusive breastfeeding duration and observed improved precision in the ATT for maternal ADHD. Paternal ADHD and 6-month exclusive breastfeeding were associated with a smaller effect size than for maternal ADHD. Men’s ability to self-regulate and respond to their partner’s breastfeeding needs could plausibly influence maternal continuation or discontinuation of exclusive breastfeeding. This finding is consistent with a qualitative review of fathers’ perceptions of infant feeding (N=20 studies), in which Baker et al concluded that men facilitate the continuation or discontinuation of breastfeeding through advocacy and practical/emotional support. However the review by Baker et al also summarized cases of domestic abuse in the Canadian setting, in which coercive methods had been used when fathers’ and mothers’ choices differed regarding infant feeding methods. We observed a small effect size for paternal ADHD on IPV at 3 months (Cohen *d* - 0.30), suggesting coercion is a plausible mechanism.

Our findings indicate that the association between maternal ADHD and 6-month exclusive breastfeeding may be confounded by maternal childhood adversity. Childhood maltreatment consistently predicts more severe ADHD symptoms and diagnosis [29,30]. Childhood adversity is also associated with life stress and plausibly related to difficulties with persistence in exclusive breastfeeding [31]. This study is the first to account for confounding by maternal childhood adversity when estimating the causal effect of maternal ADHD on 6-month exclusive breastfeeding.

### Strengths and Limitations

In this study, the breastfeeding measure may not reflect continuous exclusive breastfeeding duration, as mothers were asked to recall their feeding method over the last 7 days only. This could lead to non-differential misclassification of the outcome, biasing estimates towards the null. However 6-month exclusive breastfeeding in our sample was reported among 28-38% of mothers, which is consistent with Canadian population estimates over the same time period (34%) [32].

The ASRS is reported to have high positive predictive value [17]. However this instrument captures only externalized hyperactivity symptoms – not internalized ones – therefore participants’ exposure group may be misclassified. Moreover, girls/women in particular display hyperactivity and other externalizing symptoms to a lesser extent than boys/men, and display more internalizing symptoms such as anxiety and depression [33,34]. Self-report questionnaires and resulting non-differential misclassification of the exposure may explain the observed causal effect estimates as the ATT is likely biased towards the null.

Causal effect estimates require greater statistical power to detect a statistically significant effect. We observed a plausible, statistically significant negative effect of maternal ADHD on 6-month exclusive breastfeeding among families with maternal ADHD, a clinically relevant therapeutic target for promoting infant development.

No genetic variants are known to influence both parental ADHD risk and exclusive breastfeeding. Moreover, breastfeeding is a personal choice that depends on maternal ability [28]. Given ADHD’s strong genetic component, Mendelian randomization analyses could further clarify the causal effect of maternal ADHD on 6-month exclusive breastfeeding [20]. The exposure would then be ‘genetically predicted ADHD’, isolating the component unaffected by residual confounding; effect estimation would thus require a very large sample size.

### Generalizability

First, for ethical reasons, the IMPACT study excluded women with major psychiatric conditions including bipolar disorder, psychosis, and major depressive disorder, as well as those experiencing suicidality [15]. However, among women with ADHD, major psychiatric comorbidities such as major depressive disorder can lead to difficulties with breastfeeding [3,35]. A mixed-methods review of infant feeding practices and support among women with major psychiatric disorders (N=18 studies) concluded that these women face unique barriers to initiating and maintaining breastfeeding, including inadequate advice and infant feeding support from primary care providers and difficult choices about feeding methods and the use of psychotropic medication [14]. Our findings are thus based on a study population that excludes women at highest risk of parenting challenges, including infant feeding. We would expect the estimated causal effect of maternal ADHD on 6-month exclusive breastfeeding to be greater in women with major psychiatric comorbidities, but this is best analyzed in a separate study given the different mechanisms involved. Second, IMPACT participants also have higher socioeconomic status than the general Canadian population – particularly in educational attainment and employment during pregnancy – which could have attenuated ATT estimates [15]. Notably, according to Statistics Canada, in 2021 68% of Canadian women aged 25–64 had completed college or university, and proportions are increasing [36]. Third, the IMPACT study focused only on mother-father dyads, meaning our findings may not generalize to all family compositions (eg, same-sex couples, single-parent households). Replicating this study in an administrative health data-based cohort could help validate our findings, as the results would come from a larger, more representative sample, including participants who are at higher risk of discontinuing exclusive breastfeeding within 6 months.

Future evaluations aiming to support informed feeding decisions and maternal well-being should be mother-centered [28]. Mothers experiencing ADHD-related challenges may benefit from additional anticipatory guidance, simplified written feeding plans, reminders, structured follow-up, assistance with organizing feeding routines, and timely access to lactation consultants.

Breastfeeding support should be individualized, nonjudgmental, respectful of maternal feeding preferences, and include assessments of breastfeeding self-efficacy, a known effect modifier of exclusive breastfeeding [37].

### Conclusions

This study is the first to estimate the causal effect of maternal ADHD on exclusive breastfeeding through 6 months postpartum. Among 3217 mother-father dyads from a prospective cohort study based in Canada (2014-2020), we analyzed 6-month exclusive breastfeeding as a function of maternal ADHD, defined using a psychometrically validated scale. Among families with maternal ADHD, women’s intention to breastfeed exclusively was almost universal, yet we observed lower 6-month exclusive breastfeeding with maternal ADHD (average treatment on the treated). Our findings suggest that individualized, nonjudgmental breastfeeding support recognizing maternal neurodevelopmental differences is critically needed. Pediatric and primary care providers can be trained to identify feeding difficulties and connect families with more accessible supports to help reach therapeutic infant feeding targets.

## Supporting information

Multimedia Appendix 1

Multimedia Appendix 2

## Data Availability

Raw deidentified data from the IMPACT study are available upon reasonable request to the senior author (CLD) for planned analyses, with permission from the research team.

## END STATEMENTS

### Author Contributions

**TD:** Conceptualization, Data curation, Formal analysis, Funding acquisition, Investigation, Methodology, Software, Validation, Visualization, Writing – original draft, Writing – review & editing. **CLD:** Resources, Writing – review & editing.

