## Supplementary material for "Maternal ADHD and exclusive breastfeeding: A prospective community-based cohort study": Multimedia Appendix 1

### eMethods 1

**Variables.**
Y: outcome, exclusive breastfeeding until 6 months (binary, 1 = yes)
A: exposure, maternal ADHD (binary, 1 = yes)
L: confounder centered on its mean value in the maternal ADHD subgroup (continuous)
$\mu$: intercept
$\epsilon$: error term.

**Linear Probability Model.**
For $i = 1, ..., n$, where $n$ is the number of units (ie, mother-father dyads),

$$Y_i = \mu + \beta_1 A_i + \beta_2 L_1 i + \beta_1 \beta_2 A_i L_1 i + \epsilon_i \tag{1}$$

**Average Treatment Effect on the Treated.**
In this case, the causal estimand (ATT) corresponds to the regression model estimate,

$$ATT = \hat{\beta}_1$$

with heteroskedasticity-robust standard error estimated via the sandwich method and 95% confidence intervals estimated using the bootstrap.
