## Supplementary material for "Maternal ADHD and exclusive breastfeeding: A prospective community-based cohort study": Multimedia Appendix 2

**eTable 1.** Participants' baseline characteristics by complete-case status.

| | Complete-cases[a] | | | | | |
|---|---|---|---|---|---|---|
| | **Yes** | | **No** | | **SMD[b]** | |
| | **Mothers (n=2071)** | **Fathers (n=2071)** | **Mothers (n=1146)** | **Fathers (n=1146)** | **Mothers** | **Fathers** |
| **Socio-demographics** | | | | | | |
| Canadian born, No. (%) | 1686 (81.5) | 1611 (77.8) | 843 (73.9) | 807 (71.0) | 0.182 | 0.155 |
| Newcomer, No. (%) | 70 (18.5) | 99 (21.9) | 72 (24.7) | 74 (23.2) | 0.150 | 0.032 |
| Unemployment, No. (%) | 196 (9.5) | † | 130 (11.4) | † | 0.063 | 0.095 |
| Childhood adversity, number of ACE[c], median [IQR[d]] | 3.0 [2.0, 5.0] | 4.0 [2.0, 5.0] | 4.0 [2.0, 5.0] | 4.0 [2.0, 6.0] | 0.105 | 0.056 |
| **Substance use, pregnancy** | | | | | | |
| Cigarettes, No. (%) | † | 200 (9.7) | 63 (5.5) | 181 (15.9) | 0.171 | 0.187 |
| Alcohol, No. (%) | 340 (16.4) | 1630 (78.7) | 144 (12.6) | 807 (70.7) | 0.109 | 0.186 |
| Other substances, No. (%) | † | 156 (7.5) | † | 81 (7.1) | 0.079 | 0.017 |
| **Mental health, pregnancy** | | | | | | |
| Depression, No. (%) | 185 (8.9) | 102 (4.9) | 127 (11.1) | 58 (5.1) | 0.072 | 0.007 |
| Anxiety, No. (%) | 300 (14.5) | 200 (9.7) | 194 (17.0) | 124 (10.8) | 0.068 | 0.039 |
| **Infant characteristics** | | | | | | |
| Sex at birth, female, No. (%) | 1045 (50.5) | | 520 (45.5) | | 0.099 | |
| Preterm birth, No. (%) | † | | † | | 0.098 | |
| Low birth weight, No. (%) | † | | † | | 0.011 | |
| No NICU[e] use, No. (%) | 1914 (92.5) | | 1013 (88.8) | | 0.127 | |
| **Exclusive breastfeeding** | | | | | | |
| No intention, No. (%) | † | 380 (19.4) | 222 (19.4) | 402 (40.6) | **0.540** | **0.477** |

**[a]: Complete-cases for the dataset including the following variables: maternal childhood adversity, maternal ADHD, paternal ADHD, and 6-month exclusive breastfeeding.**

**[b]: Standardized mean difference, SMD. Significant differences are >.2 and highlighted in bold.**

**[c]: Adverse Childhood Experiences International Questionnaire (ACE-IQ) [1].**

**[d]: Interquartile range, IQR.**

**[e]: Neonatal Intensive Care Unit, NICU.**

**†: Relative frequency <5%.**
